# Evaluating the Effectiveness of International Travel Controls to Identify Monkeypox Virus Infected Travelers

**DOI:** 10.1101/2025.01.10.25320302

**Authors:** Keisuke Ejima, Yuqian Wang, Akira Endo, Hiroaki Murayama, Yun Shan Goh, Alex R. Cook, Yong Dam Jeong, Shingo Iwami, Hyeongki Park, Borame Sue Lee Dickens, Shihui Jin, Jue Tao Lim, Conrad En Zuo Chan, Po Ying Chia, Barnaby E. Young, Yang Yang, Martin Chio, David Chien Lye, Marco Ajelli

## Abstract

**Introduction:** In August 2024, the World Health Organization (WHO) declared a public health emergency due to the rapid spread of mpox in African and beyond. International travel controls (ITCs), such as health screening and viral testing, could help avoid/delay the global spread of the monkeypox virus (MPXV), fostering preparedness and response efforts. However, it is not clear whether the viral tests at immigration are sufficient to avoid importation of MPXV and which samples should be used on the viral tests.

**Methods:** We conducted a simulation study using epidemiological and viral load data to assess the effectiveness of health screening and PCR testing at immigration. This provides estimates of the proportion of infected travelers identified with this policy. Viral dynamics models were used to estimate false-negative rates of PCR tests with different detection limits according to testing regimens at three different sites: oropharynx, saliva, and rectum. We also simulated the effects of these border control methods on the recommended duration of a monitoring period for travelers from mpox-affected regions, during which individuals would self-monitor for symptoms and practice cautionary behavior.

**Results:** Our results show that the combination of health screening and PCR testing of saliva swabs under an endemic scenario identify only 74% of MPXV infected travelers. The use of rectal swabs combined with health screening allows the identification of a marginally larger share of infected travelers (79%) compared to saliva swabs. A similar identification rate could be achieved by using more sensitive PCR tests (detection limit [DL]: 10 copies/mL vs. 250 copies/mL used in our baseline analysis). We estimated that travelers from mpox-affected areas should monitor themselves and practice precautionary behavior for 16 days.

**Conclusion:** Health screening and PCR testing at immigration are likely to miss a significant proportion of MPXV-infected travelers, thus a lengthy quarantine period would be required to prevent onward local transmission. Careful consideration on other factors such as economic costs and likelihood of widespread local outbreak will need to be weighed against the adoption of these measures to prevent local mpox transmission given MPXV transmissibility and severity.

## Introduction

Monkeypox virus (MPXV), the causative agent of mpox, has caused sporadic infections in African countries for over 50 years mainly due to zoonotic spillover^1^. Two clades, I and II, have been observed in Central and West Africa, respectively^2^. Clade I is associated with more severe symptoms and higher mortality rates^2^. In 2022, human-to-human transmission, mainly during sexual intercourse, of clade IIb caused a global epidemic^3^. In September 2023, a different subtype of mpox (clade Ib) began to spread in the Democratic Republic of Congo (DRC). Notably, unlike the clade IIb epidemic, which predominantly impacted men who have sex with men (MSM) populations, clade Ib infection was reported in a substantial number of children and commercial sex workers^4,5^, raising significant public health concerns. In response, the World Health Organization (WHO) declared the mpox outbreak a public health emergency of international concern^6^. Furthermore, importations of the same subtype were identified in non-African countries, including Thailand, Sweden, India, Germany, the United Kingdom, the United States, Canada and Belgium. Moreover, secondary transmission was also found in the United Kingdom and Germany^7^, raising concerns about its potential for further international spread.

Implementing effective international travel controls (ITCs) is instrumental to reduce the spread of infections to other regions and is especially important when local transmission is not yet established. Common ITCs include health screenings, viral testing (e.g., PCR and antigen tests), case isolation, and quarantine and/or active monitoring of suspected cases^8^. Although avoiding case importation entirely may not be feasible, delaying the spread may gain time to prepare resources and plan strategies to mitigate the impact of potential epidemics. In particular, health screening and viral tests play a critical role in preventing the importation of infected individuals. Health screening aims to detect infected individuals who are symptomatic, while viral tests are used both to confirm infection in symptomatic individuals and identify pre-symptomatic and asymptomatic infected individuals. Given the relatively long incubation period of mpox (approximately 8 days both for the clade IIb strain responsible for the 2022 outbreak and for historical strains – clade Ia and IIa)^9^, the probability of individuals traveling while pre-symptomatic is non-negligible.

To delay (and/or potentially avoid) an epidemic being seeded by international travelers, health screenings and viral tests are often paired with quarantine periods for travelers^10,11^. Quarantine measures vary widely depending on the pathogen’s severity and transmissibility and the political milieu. The implementation can range from quarantine in dedicated facilities, as seen in the early stages of the COVID-19 pandemic in China and other countries with effective pandemic control, to self-quarantine at home^12^, or simply monitoring symptoms and practicing precautionary behavior^13^. Typically, the duration of quarantine is determined by the incubation period of the virus (e.g., the 95th/99th percentile). For instance, during the early phase of the COVID-19 pandemic, the U.S. Centers for Disease Control and Prevention (CDC) initially recommended a 14-day quarantine for individuals exposed to SARS-CoV-2^14^, based on epidemiological evidence suggesting that the 99th percentile of the incubation period was 14 days^15^. However, for travelers from disease-prevalent regions, such long quarantine periods may be overly conservative and suboptimal. For example, while the mean incubation period of H1N1-2009 was 2.1 days^16^, the mean time from departure to symptom onset for (infected) Japanese travelers returning from Hawaii was only 0.7 days^17^.

For mpox, several questions remain regarding international travel controls (ITCs) and quarantine measures. First, it is uncertain whether viral testing bundled together with health screening at immigration is sufficient to prevent the importation of MPXV. Second, with multiple sampling sites available, including saliva and rectum (details provided in the next section), it is unclear which sample type should be prioritized for viral testing. Third, if viral testing proves insufficient, the appropriate quarantine duration for travellers from mpox-affected countries who may have been exposed remains to be determined. To address these questions, we developed mathematical models informed by viral load data and epidemiological insights.

## Methods

### False negative rates from viral dynamics model

Critical to determining the effectiveness of border measures is the false negative rate of viral tests, including PCR tests, which is influenced by the viral load and the ability of the test to detect the virus (detection limit). Since the viral load varies throughout the course of an infection, increasing initially, reaching a peak, and then declining, the false negative rate also changes over time^15^. In a previous study, we estimated false negative rates for PCR tests for SARS-CoV-2 using a mathematical model of within-host viral dynamics, considering the timing of tests and different detection limits^18^. Here we adapted the same approach to estimate the false-negative rate of PCR tests for MPXV.

#### MPXV viral load data

To parameterize the viral dynamics model, we analyzed two prospective longitudinal cohort studies. The first cohort enrolled 77 men with acute mpox infections (clade IIb) hospitalized in Shenzhen, China, between June 9 and November 5, 2023^19^. Participants were followed up to 21 days after symptom onset. Samples were collected every 2-3 days from various sites, including skin lesions, rectum, saliva, oropharynx, urine, and plasma. The second cohort enrolled 25 individuals who had high-risk contact with mpox infected patients in Antwerp, Belgium, between June 24 and July 31, 2022^20^. Participants were followed up for a maximum of 21 ± 2 days after inclusion, while infected participants were monitored until they tested MPXV-PCR positive with typical mpox symptom onset and then referred to routine clinical care. During weekly follow-up visits, blood, saliva, oropharyngeal swabs, genital swabs, anorectal swabs, and skin lesion swabs were collected. Additionally, anorectal swabs, genital swabs, and saliva were also collected through daily self-sampling. PCR sensitivity was higher with oropharynx, saliva, rectum, and skin lesions (>70%), while PCR sensitivity was lower with urine, serum, genital swabs and plasma samples (<50%). Hence, we have focused our analysis on samples with higher PCR sensitivity (oropharynx, saliva, and rectum) for ITC purposes. It is worth noting that we have excluded the skin lesion data in the analysis, as PCR testing on skin lesions can only be conducted after lesions are visible. Therefore, relying on skin lesions for PCR testing at immigration to identify infected individuals is moot. Quantitative reverse transcription PCR (qRT-PCR) was used to measure cycle threshold (Ct) values. To convert the Ct value to viral load, we applied the following equation: 𝐶𝑡 = −3.611(𝑙𝑜𝑔_10_ 𝑅𝑁𝐴) + 41.388 as described in the paper^19^. Ct values > 40 (i.e., < 10^3^ copies/mL for viral load) are considered negative.

#### Viral dynamics model and fitting

To characterize the MPVX viral dynamics, we used the following system of ordinary differential equations, which accounts for the fundamental biological process of the infection within a host, including viral replication and elimination due to immune response:

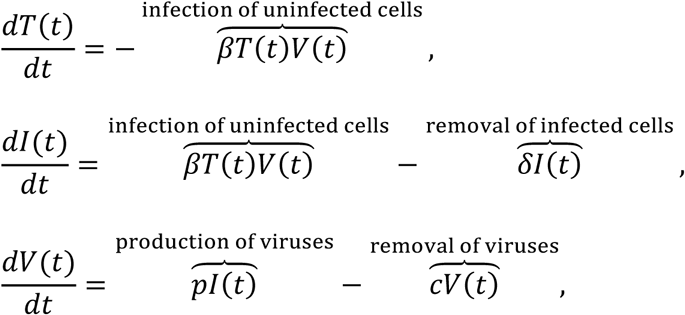

where the variables 𝑇(𝑡), 𝐼(𝑡), and 𝑉(𝑡) represent the number of uninfected target cells, number of infected target cells, and amount of virus at 𝑡 days after symptom onset, respectively. These represent the infection of uninfected cells at rate 𝛽𝑇(𝑡)𝑉(𝑡) and the removal of infected cells at rate 𝛿𝐼(𝑡) cells per day, and the production of virus at rate 𝑝𝐼(𝑡) and the removal of virus at rate 𝑐𝑉(𝑡) viral units per day. Note that we use days after symptom onset as the time scale. The model parameters 𝛽, 𝛿, 𝑝, and 𝑐 represent the rate of virus infection, death rate of infected cells, (per cell) viral production rate, and (per capita) clearance rate of the virus, respectively. Under the quasi-steady-state assumption, the model is reduced to a two-dimensional model as follows:

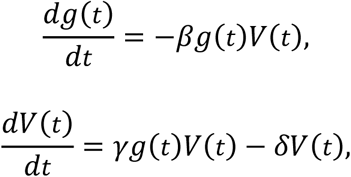

where 𝑔(𝑡) is the fraction of uninfected target cell population at day 𝑡 to that at day 0 (i.e., 𝑔(0) = 1), and 𝑉(𝑡) is the amount of virus at day 𝑡 (copies/mL), respectively. Note that time 0 corresponds to the day of symptom onset. Details on the transformation of this model are reported in the reference^21^. The quasi-steady-state assumption is generally reasonable for most viruses causing acute infectious disease because the clearance rate of virus (𝑐) is typically much larger than the death rate of the infected cells (𝛿) as evidenced by *in vivo* observations^21–23^. Thus, we estimated the four parameters: 𝛽, 𝛾, 𝛿, and 𝑉(0) (viral load at the day of symptom onset). We fit the viral dynamics model to the viral load data collected from the three different sites (rectum, saliva, and oropharynx) independently using a non-linear mixed effect model.

The nonlinear mixed-effects model incorporates fixed effects and random effects accounting for inter-site variability in viral dynamics. Specifically, the parameter for site 𝑘, 𝜃_k_(= 𝜃 × 𝑒^nk^), is represented by the fixed effect, 𝜃, and the random effect, 𝜋_k_, is assumed to follow the Gaussian distribution with mean 0 and standard deviation Ω. The fixed effect (population parameter) and random effect were estimated by using the stochastic approximation expectation-maximization (EM) algorithm and empirical Bayes’ method, respectively. We chose these methods because the EM algorithm is well-suited for estimating population-level parameters in the presence of latent variables, whereas the empirical Bayes allows us to efficiently estimate inter-site-level variations by using prior information. Using estimated parameters and a Markov Chain Monte Carlo algorithm, we obtained the conditional distribution of model parameters for each site. Left censoring was considered based on the lower limit of detection (Ct values < 40). Monolix 2023R1 (https://www.lixoft.com) was used for viral dynamics model fitting.

#### Simulation analysis to estimate false negative rates over time

We repeatedly randomly resampled a parameter set for individual 𝑖 at site 𝑘 (i.e., 𝛽_ik_, 𝛾_ik_, 𝛿_ik_, and 𝑉(0)_ik_) from the estimated posterior distributions, and ran the viral dynamics model. The viral load obtained by running the viral dynamics model is considered as the “*true” viral load*, 𝑉_ik_(𝑡). However, PCR test results are influenced by measurement error. To account for this, we added measurement error to the true viral load to simulate *measured viral load*, 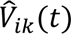, whereby 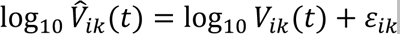, 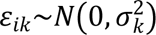^24,25^. As the time scale of the viral dynamics model is days after symptom onset, we corrected the time scale to time after infection by adding incubation period, 𝜂, drawn from the incubation period distribution obtained from the meta-analysis^9^. That says, 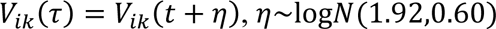 (mean=8.1, 95th percentile=18), where 𝜏 is the infection age (i.e., the time which has elapsed since infection). The error distribution is obtained by fitting a normal distribution to the residuals (i.e., the difference between the common logarithms of the true viral load and the measured viral load). We repeated this process 10,000 times to create the viral-load distribution over time. The false-negative rate at site 𝑘 at infection age 𝜏 is computed as the proportion of cases with a viral load below the detection limit: 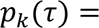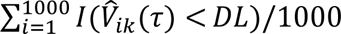, where 𝐷𝐿 is the detection limit, and 𝐼 is the identity function. We used 250 copies/mL as the detection limit of PCR tests^26^. As a sensitivity analysis, we performed the same simulation using different detection limits, namely 10 and 1,000 copies/mL.

### Detection rates by health screening and PCR tests and post-entry incubation periods for travelers

To assess the impact of health screenings and PCR tests, we examined a population of individuals infected during travel in mpox-affected countries and moving to non-mpox-affected countries. Upon arrival in non-mpox-affected countries, the time elapsed since infection, referred to as the infection age (𝜏), represents the duration between the date of infection and the date of arrival at the travel destination. Assuming that the mpox cases still were exponentially increasing at the country of origin at a growth rate 𝑟 and travelers are exposed proportionally to the number of the cases, the number of infected (traveling) individuals with infection age 𝜏 at immigration is described as follows: 𝑗(𝜏) = 𝑗_0_exp (−𝑟𝜏), where 𝑗_0_ is a constant number representing those at infection age 𝜏 = 0 (𝑗(0) = 𝑗_0_). Given that health screenings at immigration detect individuals with symptoms, we aim to use PCR tests to identify asymptomatic individuals at the time of entry. These individuals are modeled as being in the early stages of infection, where the infection age 𝜏 is below the threshold for symptom manifestation. The probability of detecting an asymptomatic individual with a PCR test is described as a function of viral load dynamics over 𝜏, modeled as 𝑗_pre_(𝜏) = 𝑗_0_exp (−𝑟𝜏)𝐿(𝜏), where 𝐿(𝜏) is the survival function of symptom development by infection age 𝜏. The survival function of symptom development 𝐿(𝜏) can be computed from the probability density function of incubation period, 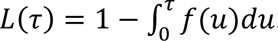. The number of pre-symptomatic individuals who test negative for mpox on PCR tests conducted at immigration is represented by 𝑗_pre,neg_(𝜏) = 𝑗_0_exp (−𝑟𝜏)𝐿(𝜏)𝑝(𝜏), where 𝑝(𝜏) is the false-negative rate of PCR tests at infection age 𝜏. The proportion of infected travelers who develop symptoms and are subsequently identified by the health screenings at immigration (including those who cancelled travel due to symptoms) can be modeled as 𝐻 = 1 − 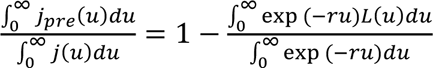. This assumes that the health screenings detect all symptomatic mpox cases. The proportion of infected travelers identified through PCR testing can be modeled as 𝑇 = 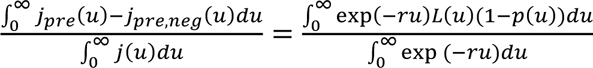. Note that 𝑗_0_ is cancelled in the derivation process of 𝐻 and 𝑇.

The quarantine period can be determined based on the incubation period, assuming that the time of exposure is known – typically the 95^th^ percentile of the incubation period distribution^27^. However, in this study, we assume that the timing of exposure is not known and only the timing of entrance to the country is known. Therefore, the quarantine period begins upon entry into the country. To determine the appropriate length, we estimated the remaining incubation period for pre-symptomatic individuals at immigration (time from immigration to symptom onset, hereafter referred to as *post-entry incubation period*). The 95th, 80th, and 70th percentiles of this post-entry incubation period were used to define the quarantine period. We emphasize here that quarantine refers to self-monitoring for mpox symptoms and practicing precautionary behavior (e.g., abstinence).

From the mathematical standpoint, we set the time after immigration, 𝑠, as a time scale. Given a maximum travel duration of 𝑘 days, the number of individuals that developed symptoms at time 𝑠, 𝑖(𝑠), can be modeled as

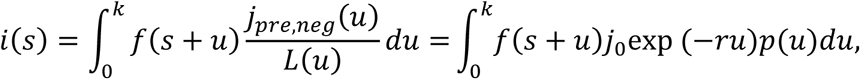

because those who are presymptomatic and test-negative on mpox PCR test at immigration with infection age 𝑢, 𝑗_pre,neg_(𝑢), have already survived (i.e., did not develop symptoms) for 𝑢 days at the time of immigration, 𝐿(𝑢), and develop symptoms 𝑠 days after immigration, when their infection age is 𝑠 + 𝑢. In other words, the distribution of a presymptomatic and test-negative infected traveler’s time from immigration to symptom onset, 𝑠, conditional to time from exposure to immigration being 𝑢 and the absence of symptoms by immigration is 𝑓(𝑠 + 𝑢)/𝐿(𝑢). As 𝑖(𝑠) is a number, it can be normalized to be a probability density function of the post-entry incubation period as 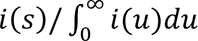. Note that 𝑗 is cancelled after normalization. We also conducted sensitivity analyses under an epidemic scenario assuming 𝑅_0_ = 1.5 ^28^. **Supplemental Table 1** summarizes the variables used in this study. All epidemiological parameters except false-negative rate (obtained by fitting the viral dynamics model to the viral load data) are obtained from literature on clade IIb except incubation period (from clade Ia, IIa, and IIb) (**Supplemental Table 2**).

## Result

### Time-and-site-dependent false-negative rates

The viral load curve for each of the four sites shows a similar trend, but remarkable differences (**Figure 1**). The peak viral load occurred at 5 days after symptom onset for rectum and oropharynx samples and 6 days for saliva samples. Peak viral load levels from the rectum samples were the highest levels (10^6.8^ copies/mL), around ten-fold higher than from saliva samples (10^5.7^ copies/mL), which in turn was around ten-fold higher then from the oropharynx samples (10^4.8^ copies/mL). MPXV remained detectable for varying durations: 19 days for rectum, 18 days for saliva, and 14 days for oropharynx. We emphasize that the length of viral shedding may exceed that of infectiousness, as the infectiousness threshold for mpox, 10^6.5^ copies/mL^29^, is higher than the detection limit. A comparison between the simulated viral load curves and the observed data for each infected individual is shown in **Supplemental Figure 1**, showing good overall concordance to the theoretical model. We also compared the estimated parameter values from the different sites (**Supplemental Figure 2**) and found a moderate correlation in the viral dynamic parameters of oropharynx and saliva. Estimated model parameters are reported in **Supplemental Table 3**.

**Figure 1:**
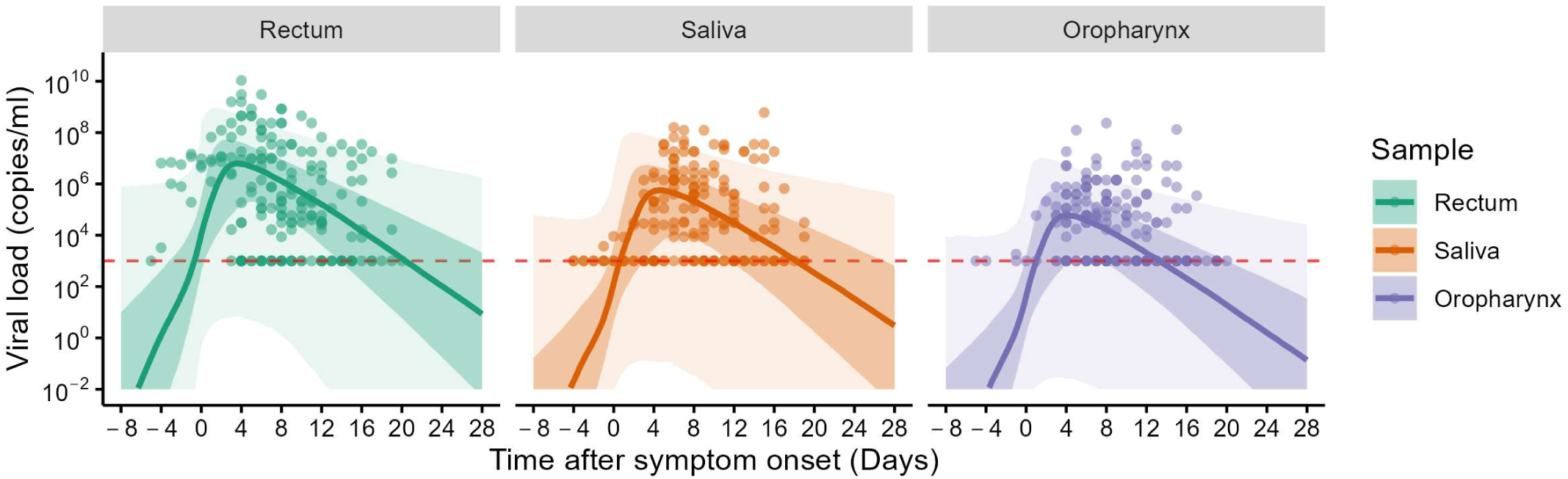
Estimated viral load over time. The thick lines are the estimated mean viral load trajectories for the three sites: rectum, saliva, and oropharynx. The shaded regions correspond to 50% and 95% CIs. The red horizontal line corresponds to the detection limit in the original study^19^ (10^3^ copies/mL).

The estimated viral dynamics allowed us to estimate false negative rate over time for various detection limits: 10, 250, and 1000 copies/mL (**Figure 2**). Using 250 copies/mL as the reference detection limit, we observed that the false negative rate is initially at 82%, 89%, 91% for rectum, saliva, and oropharynx, respectively and then decreases towards the viral load peak and subsequently increases as the viral loads fall. The lowest false negative rates were 15%, 25%, and 34% for rectum, saliva, and oropharynx, respectively, implying that even with the ideal timing, a high proportion of infections would be missed from border screening. The high false negative rate for oropharynx is explained by the relatively low mean viral load compared with saliva. Similar trends were observed for other detection limits, with higher detection limits generally associated with higher false negative rates.

**Figure 2:**
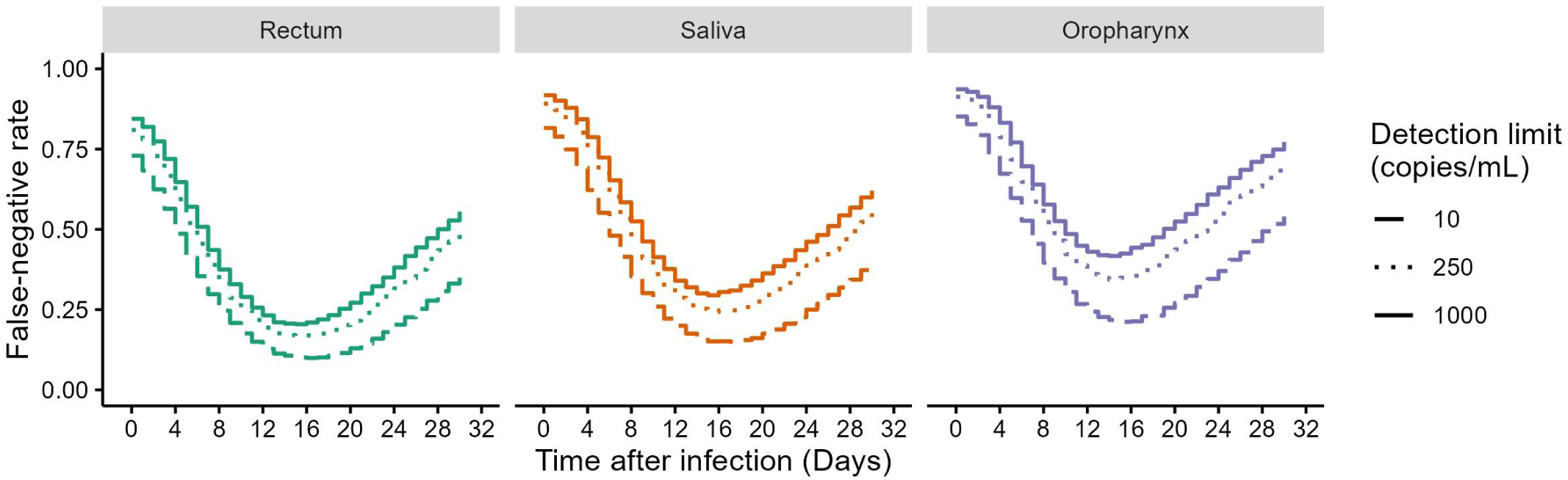
False-negative rates of PCR tests on rectum, saliva, and oropharynx The false-negative rates of PCR tests were computed for rectum, saliva, and oropharynx over time. The different types of lines correspond to different detection limits (10, 250, 1000 copies/mL).

Given the ease of saliva sampling, we selected saliva for PCR testing at immigration as a primary analysis. Rectal and oropharyngeal swabs were considered for sensitivity analyses.

### Effectiveness of health screenings and PCR tests in identifying infected travelers

Assuming an endemic situation at the source, health screenings at immigration will successfully identify 62% of infected individuals (**Figure 3**). When PCR tests are conducted on saliva samples from all pre-symptomatic individuals, an additional 12% of MPXV infected individuals will be identified (detection limit: 250 copies/mL). Assuming a more sensitive detection limit of 10 copies/mL, the detection rate would increase to 16%. A different fraction of MPXV infected travelers would be identified if rectal or oropharyngeal swabs were to be collected: 17% and 11%, respectively, assuming a detection limit of 250 copies/mL. As the mean incubation period is 8.1 days and only those who are not symptomatic could be identified by PCR tests at the border, the lower false-negative rate before the symptom onset yields to higher detection rate. Under the assumption of a growing epidemic (with 𝑅_0_ of 1.5) at the source, health screenings at immigration would identify 51% of infected individuals (**Supplemental Figure 3**). 20, 14, and 12% more cases would be identified by PCR tests on rectal, saliva, and oropharyngeal swabs, respectively.

**Figure 3:**
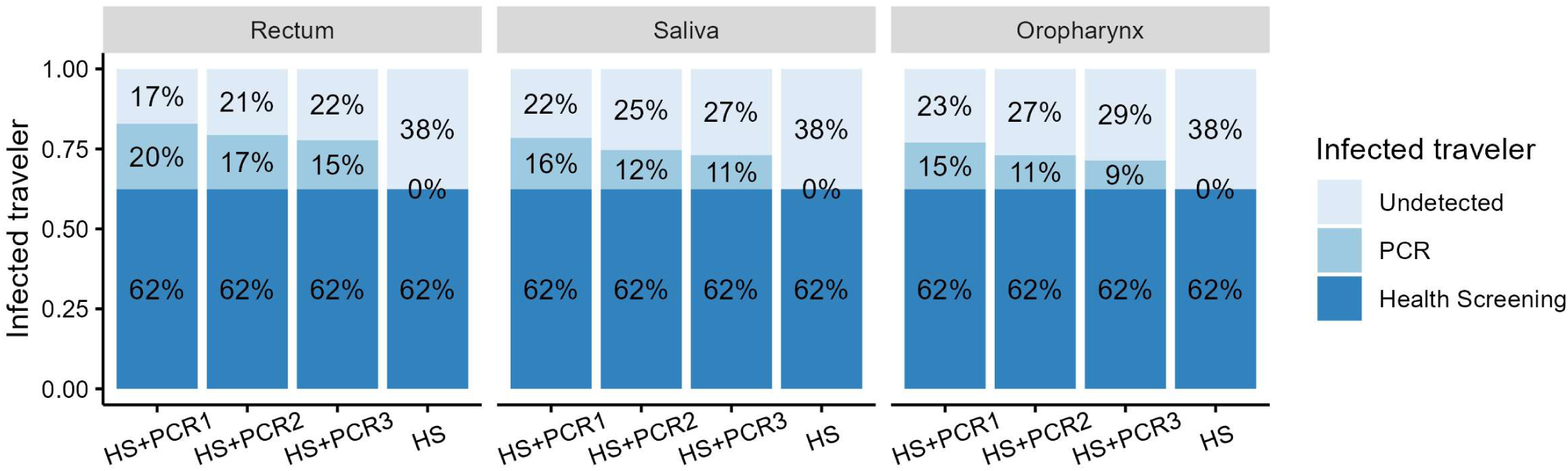
Effectiveness of health screening and PCR tests on identifying infected travelers under endemic situation. Among travelers infected in mpox-affected countries, a portion is detected through screenings based on symptom presence (dark blue) and PCR tests (light blue) conducted at immigration. We considered four scenarios: health screening only (HS), health screening combined with PCR testing at a detection limit of 10 copies/mL (HS+PCR1), 250 copies/mL (HS+PCR2), or 1000 copies/mL (HS+PCR3). For PCR testing, we simulated the use of rectum, saliva, or oropharyngeal sample assuming endemic situation.

### Length of the quarantine period

We defined quarantine periods based on the 95th, 80th, and 70th percentiles of the incubation period and post-entry incubation period distribution, as illustrated in **Figure 4**. The 95th percentile is commonly used, allowing for a 5% risk of ending quarantine before symptom development. Using lower percentiles (70th or 80th) can reduce the quarantine period, but increases the risk of infected individuals being released prematurely while still being pre-symptomatic.

**Figure 4:**
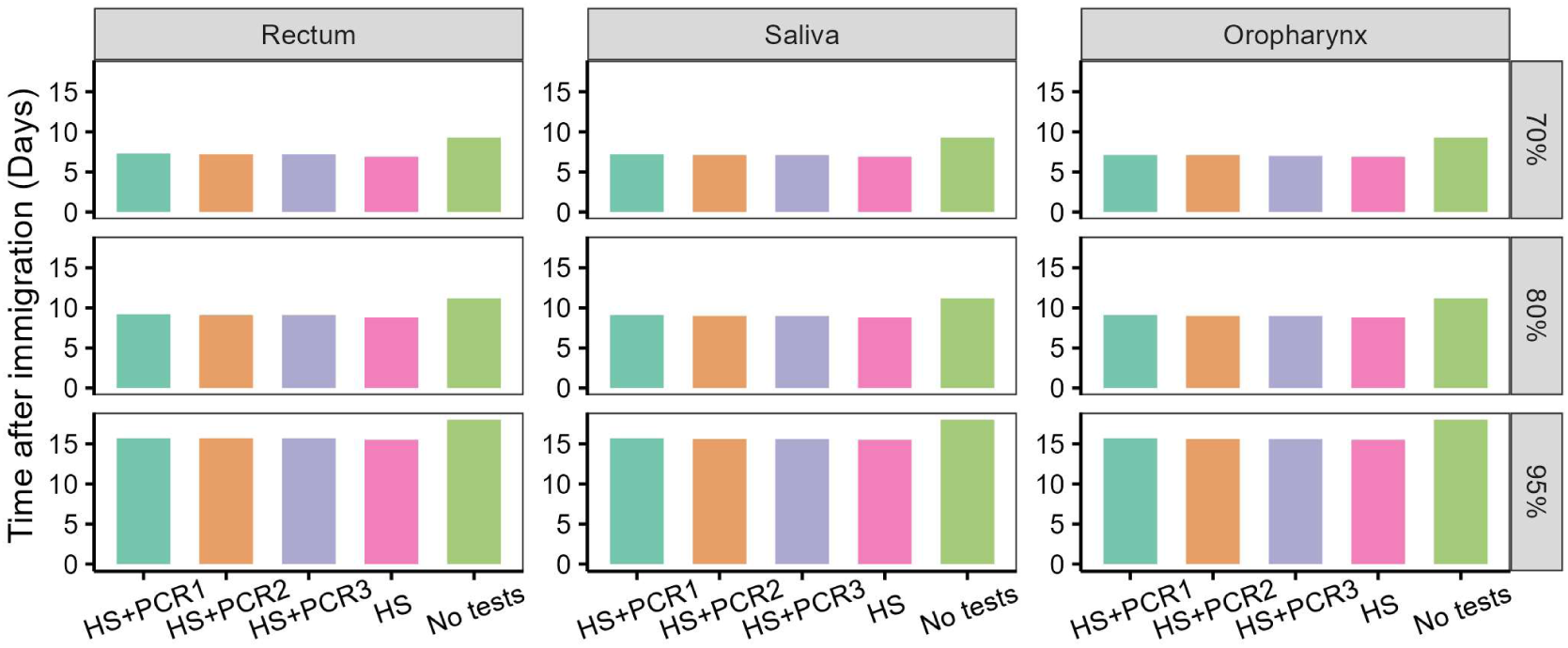
Quarantine period under health screening and PCR tests at immigration assuming endemic situation. Quarantine period was computed as 70^th^, 80^th^, and 95^th^ percentiles of (post-entry) incubation period distribution. We considered four scenarios: no health screening and PCR tests (No tests), health screening only (HS), health screening combined with PCR testing at a detection limit of 10 copies/mL (HS+PCR1), 250 copies/mL (HS+PCR2), or 1000 copies/mL (HS+PCR3) under endemic situation. For PCR testing, we simulated the use of rectum, saliva, or oropharyngeal sample assuming endemic situation.

Assuming endemicity at the source, when solely relying on the 95th percentile of the incubation period, a quarantine of 18 days is recommended. However, using the 80th or 70th percentiles reduces the quarantine period to 11 and 9 days, respectively. If health screening is implemented at immigration, the 95th percentile of the post-entry incubation period is 15.5 days, suggesting a 2.5-day reduction in quarantine period compared to using the incubation period alone. Conversely, when PCR tests (DL=250 copies/mL) are conducted, the 95th percentile of the post-entry incubation period increases slightly, suggesting a quarantine period of 15.6 days. Even when assuming epidemic situation, we confirmed the quarantine period (i.e., the 95th percentile of the incubation period) is about 15.7 days with health screening and PCR tests. **Supplemental Figure 5** illustrates the incubation period distribution and post-entry incubation period for different samples with different detection limits under epidemic and endemic situations.

Similar results were observed in sensitivity analyses that varied the percentile used to determine the quarantine period (70th and 80th percentiles) and considered different swab types for PCR testing at immigration (rectal and oropharyngeal swabs).

## Discussion

In response to the Public Health Emergency of International Concern declared by the WHO and the identification of MPXV outside the African continent, public health authorities and other major stakeholders are evaluating the possibility of implementing immigration control measures. For example, on August 23, 2024, Singapore Ministry of Health has implemented health screenings, including temperature checks and visual assessments, at international airports and seaports for travelers and crew arriving from mpox-affected regions^30^.

To evaluate the effectiveness of immigration control measures, we developed a mathematical model to assess the proportion of infected travelers identified through health screenings and PCR testing. We found that a combination of health screenings and PCR testing of saliva at immigration can identify 74% of the MPXV infected travelers. Even considering more sensitive tests (i.e., 10 copies/mL as the detection limit) or other swab sites (i.e., oropharynx and rectum samples) would not allow the identification of about 20% of MPXV infected travelers. Should public health authorities aim at fully preventing onward local transmission, combining health screening and PCR testing would need to be supplemented by a quarantine period for be required. We also provide time-varying false negative rate estimates for various detection limits, offering critical insights into the effectiveness of deploying different test types as they become available.

This study has some limitations. First, due to the limited clinical and epidemiological information available for clade Ib, most parameters were based on clade IIb, which caused the 2022 outbreak. As more relevant data becomes available for clade Ib it would be advantageous to update this modeling analysis. Second, we did not fully account for fully asymptomatic infections in our analysis. Serological surveys and screening tests have provided evidence of asymptomatic MPXV clade IIb infections^31–33^. Further research is warranted to investigate the role of asymptomatic infection in MPXV transmission. Third, we only modelled quarantine of individuals who may have been infected; therefore, we have not argued how to treat confirmed cases such as medical treatment and isolation (to avoid further transmission). Fourth, our analysis considered the time elapsed between exposure and arrival at the destination country. On one hand, this enables us to obtain more realistic estimation of the post-entry incubation period; on the other hand, this means that our findings should be revised if the MPXV affected areas start to show larger or lower rates of local transmission. Fifth, we did not consider specificity of health screening and PCR tests. In general, the specificity of PCR tests is high (and considered as the gold standard test method)^34^; however, given that mpox can present with typical symptoms (such as fever), high specificity (i.e., to minimize the risk of false positives) is important from an operational perspective in health screening.

It is important to stress that our study does not discuss whether or under which circumstances ITCs are recommended; instead, it provides quantitative estimates of the effectiveness of a combination of health screenings and PCR tests at immigration (approximately 75% of infected travelers) and of quarantine duration after arrival (approximately 2 weeks). The decision to implement public health interventions is for local authorities to make based on a wider range of considerations that go beyond the effectiveness of ITCs and quarantine, such as disease severity, likelihood of a widespread local outbreak, ability to control local spread through other measures, economic costs, strategy feasibility, among others. Furthermore, these factors may evolve over time. For example, Murayama and Asakura et al.^35^ suggested that clade Ib acquired the capacity to be sexually transmitted, which is a possible explanation for the increased outbreak potential. For this reason, careful thought should be taken on the right degree of surveillance at the border as the epidemiology changes.

## Data availability

The data that support the findings of this study are available from the corresponding authors on request.

## Code availability

All analyses were performed with the statistical computing software R (version 4.3.3) (https://www.r-project.org/). The analysis using nonlinear mixed-effects modeling was performed on Monolix 2023R1 (https://www.lixoft.com/). The study’s supporting codes can be found on Github at https://github.com/SherryYuqianWang/Mpox.

## Acknowledgments

This study was supported in part by the Ministry of Education, Singapore, under its Academic Research Fund Tier 1 Seed Award (RLMOE100201900000001), a Lee Kong Chian School of Medicine startup grant (LKCMedicine-SUG, #022487-00001), and JST, PRESTO (JPMJPR23R3) (to KE). AE is supported by Japan Society for the Promotion of Science (JP22K17329) and JST (JPMJPR22R3).

## Author contributions

Conceived and designed the study: KE, MA. Obtained and analyzed the data: KE, YW, YY, HP. Wrote the paper: KE, YW, AE, HM, YSG, ARC, YDJ, SI, HP, BSLD, SJ, JTL, CEZC, PYC, BEY, YY, MC, DCL, and MA. All authors read and approved the final manuscript.

## Supplemental Figures

**Supplemental Figure 1:**
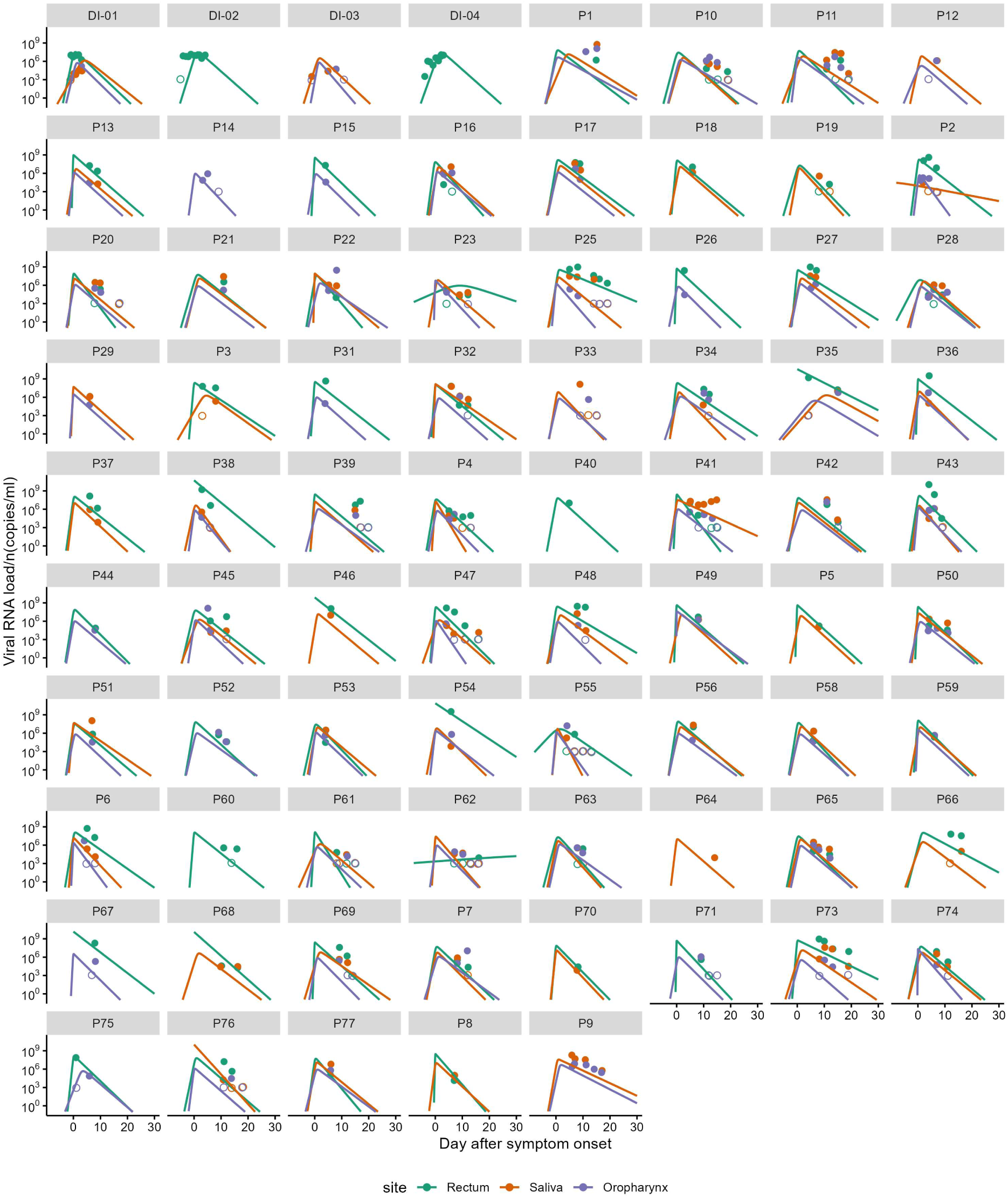
Estimated viral load curve for each infected individual The thick lines are the viral load trajectory drawn based on the best-fit parameter set for the three sites: rectum (orange), saliva (purple), and oropharynx (pink) for individuals with detectable viral load. The circles correspond to observed viral load. The viral load below the detection limit (10^3^ copies/mL) is shown by open circles.

**Supplemental Figure 2:**
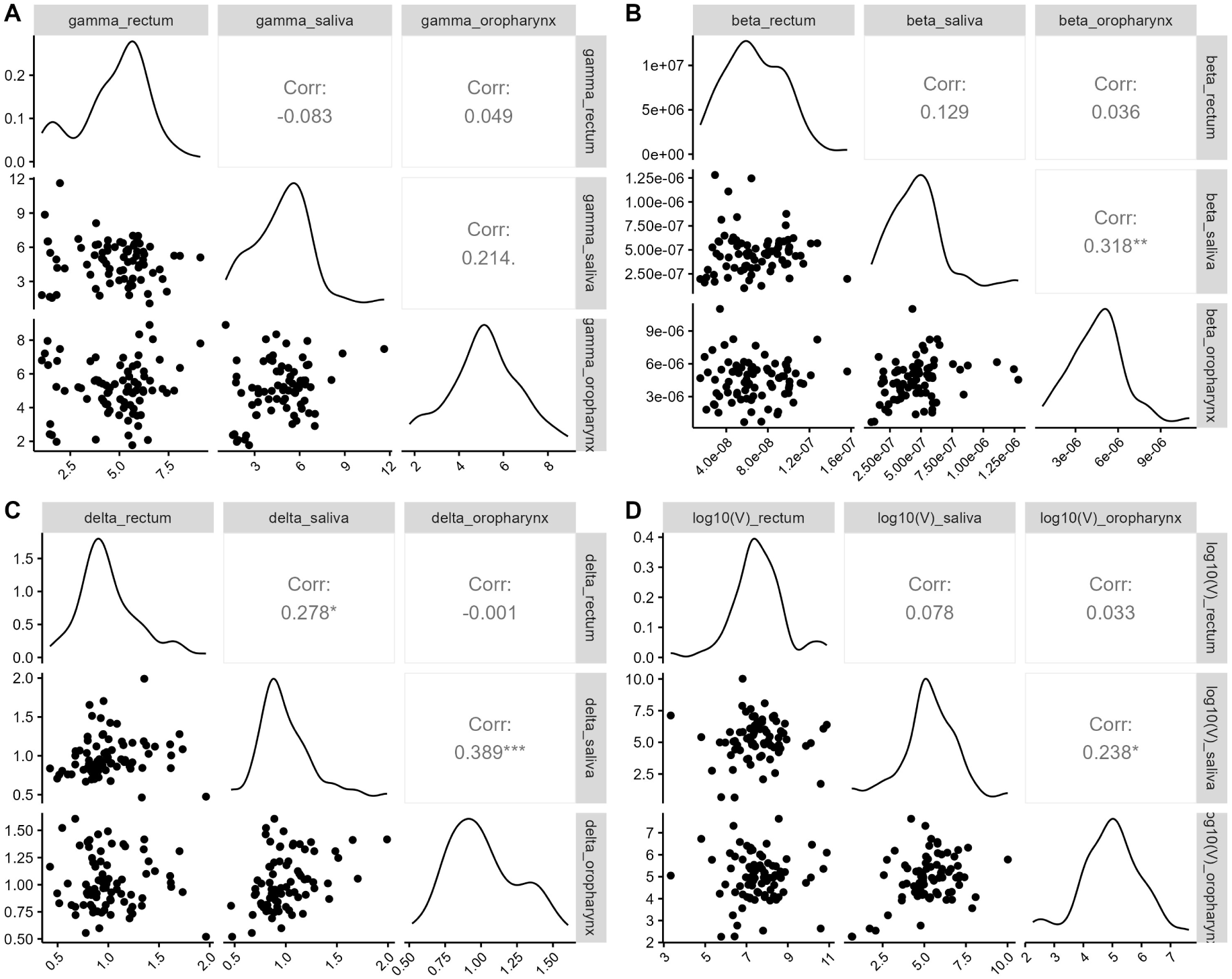
Rank correlation between individual parameter of the three sites

**Supplemental Figure 3:**
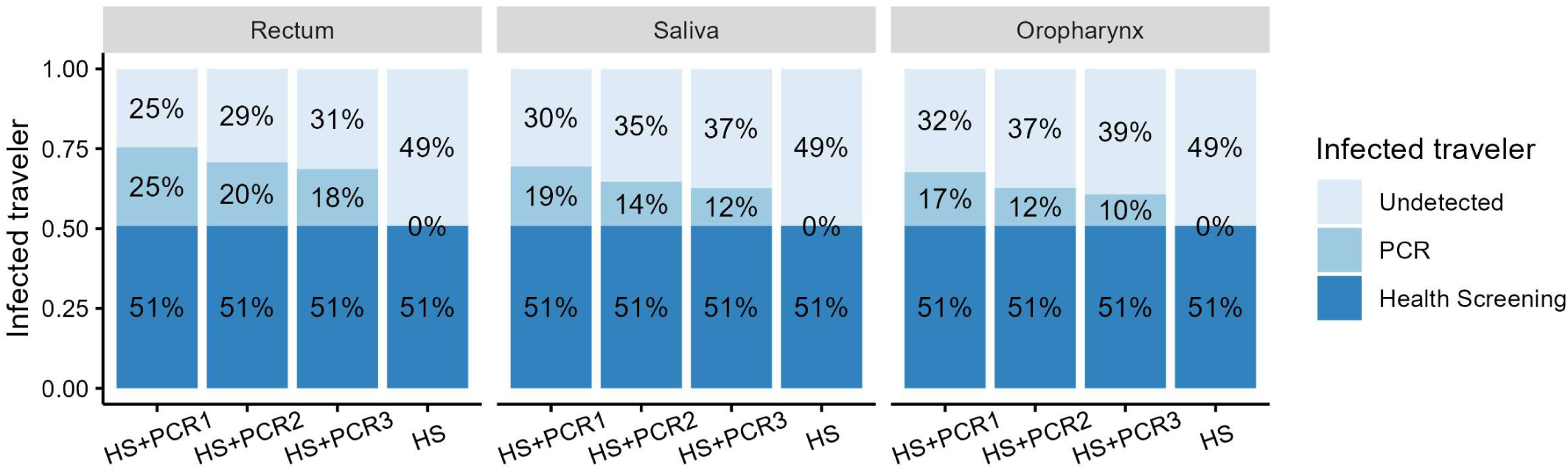
Effectiveness of health screening and PCR tests on identifying infected travelers under epidemic situation Among travelers infected in mpox-affected countries, a portion is detected through screenings based on symptom presence (dark blue) and PCR tests (light blue) conducted at immigration. We considered four scenarios: health screening only (HS), health screening combined with PCR testing at a detection limit of 10 copies/mL (HS+PCR1), 250 copies/mL (HS+PCR2), or 1000 copies/mL (HS+PCR3). For PCR testing, we simulated the use of rectum, saliva, or oropharyngeal sample under epidemic situation.

**Supplemental Figure 4:**
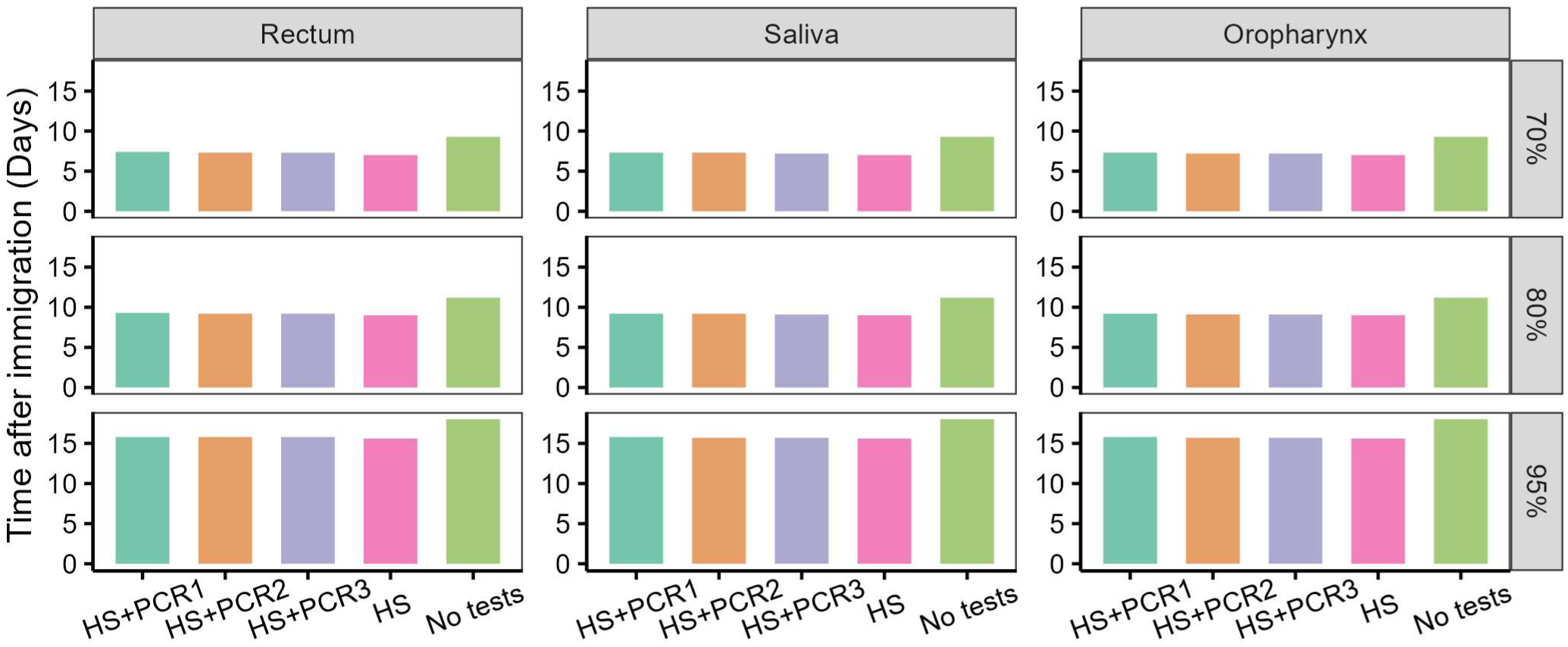
Quarantine period under health screening and PCR tests at immigration assuming epidemic situation. Quarantine period was computed as 70^th^, 80^th^, and 95^th^ percentiles of (post-entry) incubation period distribution. We considered four scenarios: no health screening and PCR tests (No tests), health screening only (HS), health screening combined with PCR testing at a detection limit of 10 copies/mL (HS+PCR1), 250 copies/mL (HS+PCR2), or 1000 copies/mL (HS+PCR3) under epidemic situation. For PCR testing, we simulated the use of rectum, saliva, or oropharyngeal sample assuming epidemic situation.

**Supplemental Figure 5:**
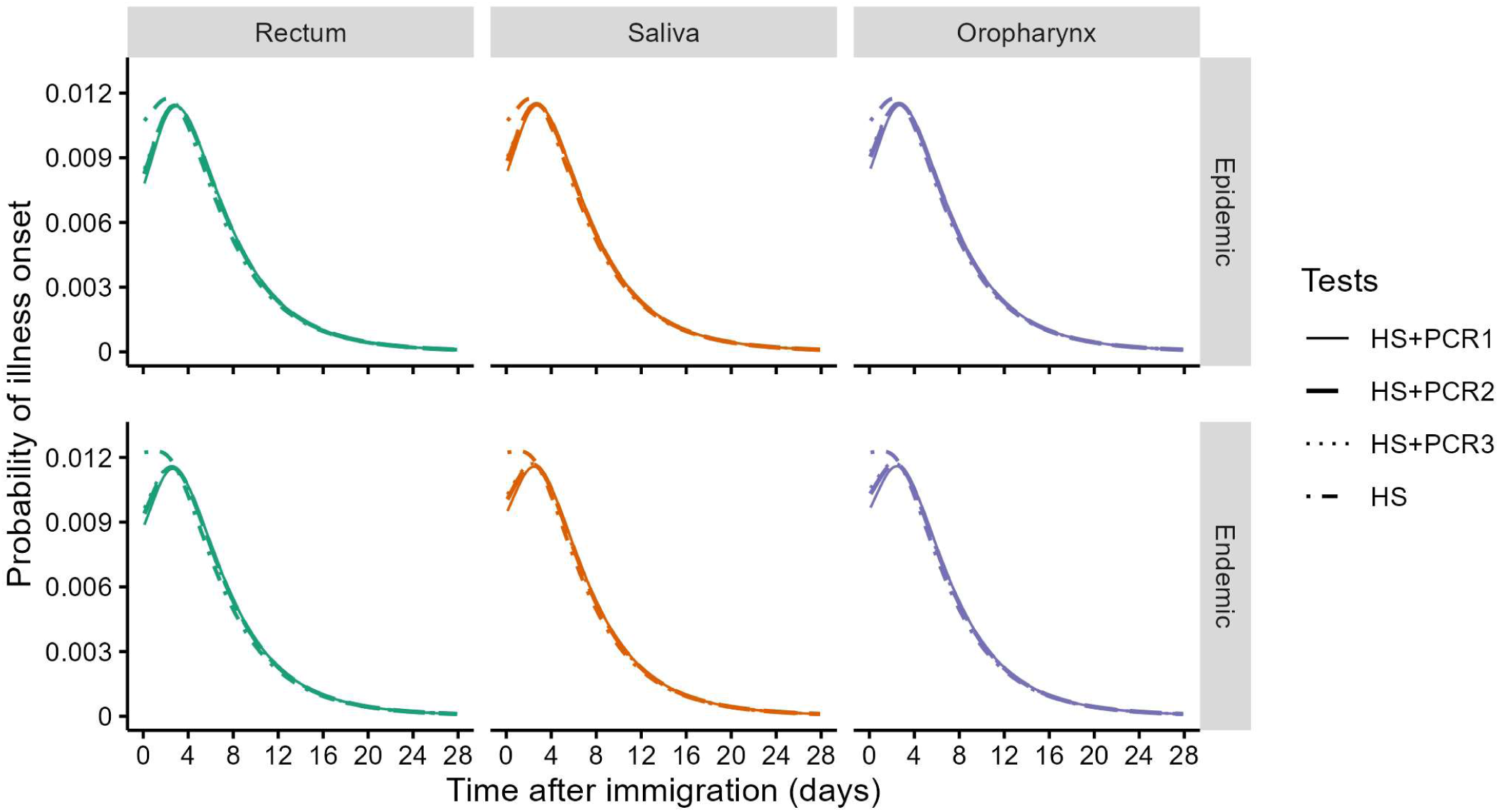
Post-entry incubation period distributions The incubation period distribution and the post-entry incubation distributions. We assumed PCR tests with different detection limits are performed at immigration (10, 250, and 1000 copies/mL). We performed the same simulation assuming PCR tests are performed on rectum, saliva, or oropharyngeal sample assuming epidemic and endemic situation.

## Supplemental Tables

**Supplemental Table 1.** Variables related to infected individuals Variables Description.

| Supplemental Table 1. Variables related to infected individuals |  |
| --- | --- |
| Variables | Description |
| $j(\tau)$ | The number of infected travelers who reach countries without mpox outbreaks at infection age, $\tau$ |
| $j_{pre}(\tau)$ | Among $j(\tau)$, the number of infected travelers who are not symptomatic (pre-symptomatic) |
| $j_{pre,neg}(\tau)$ | Among $j(\tau)$, the number of individuals who are test negative by PCR tests performed at immigration |
| $i(s)$ | The number of infected travelers who develop symptom at time $s$ |
| $p(\tau)$ | False-negative rate of PCR tests |

**Supplemental Table 2.**
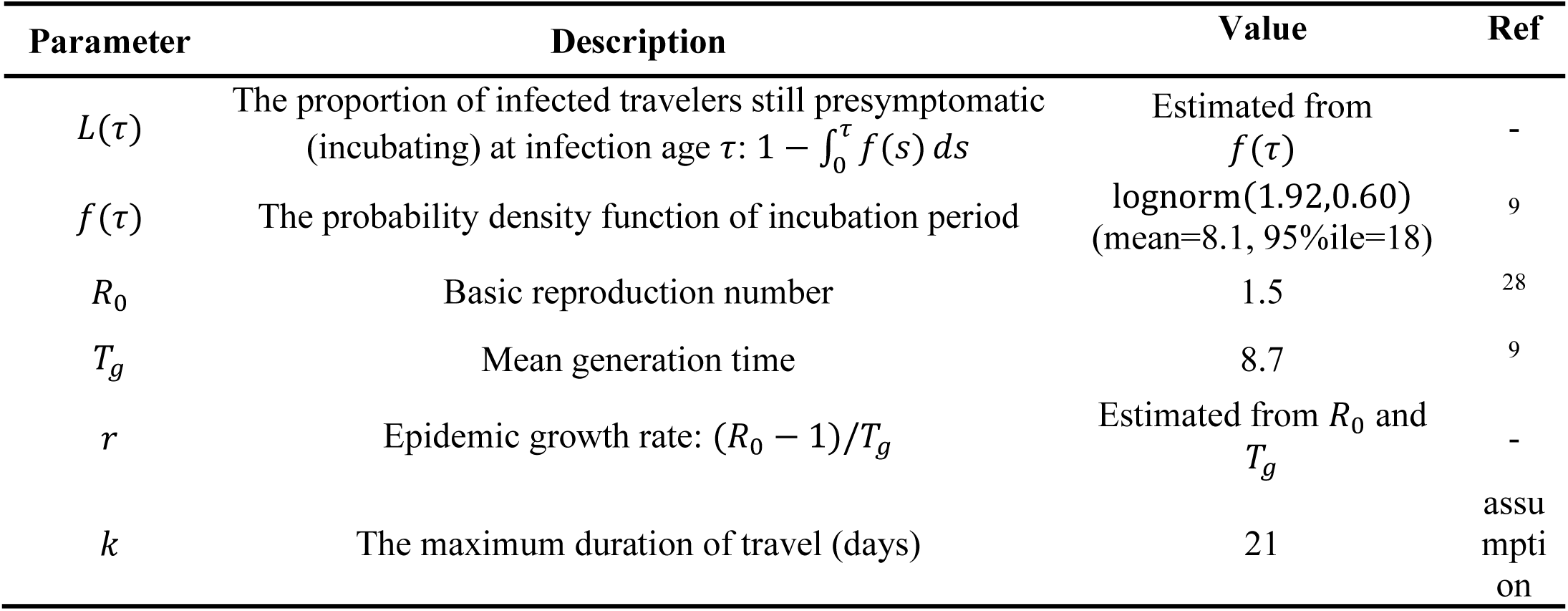
Epidemiological parameters for mpox.

**Supplemental Table 3.**
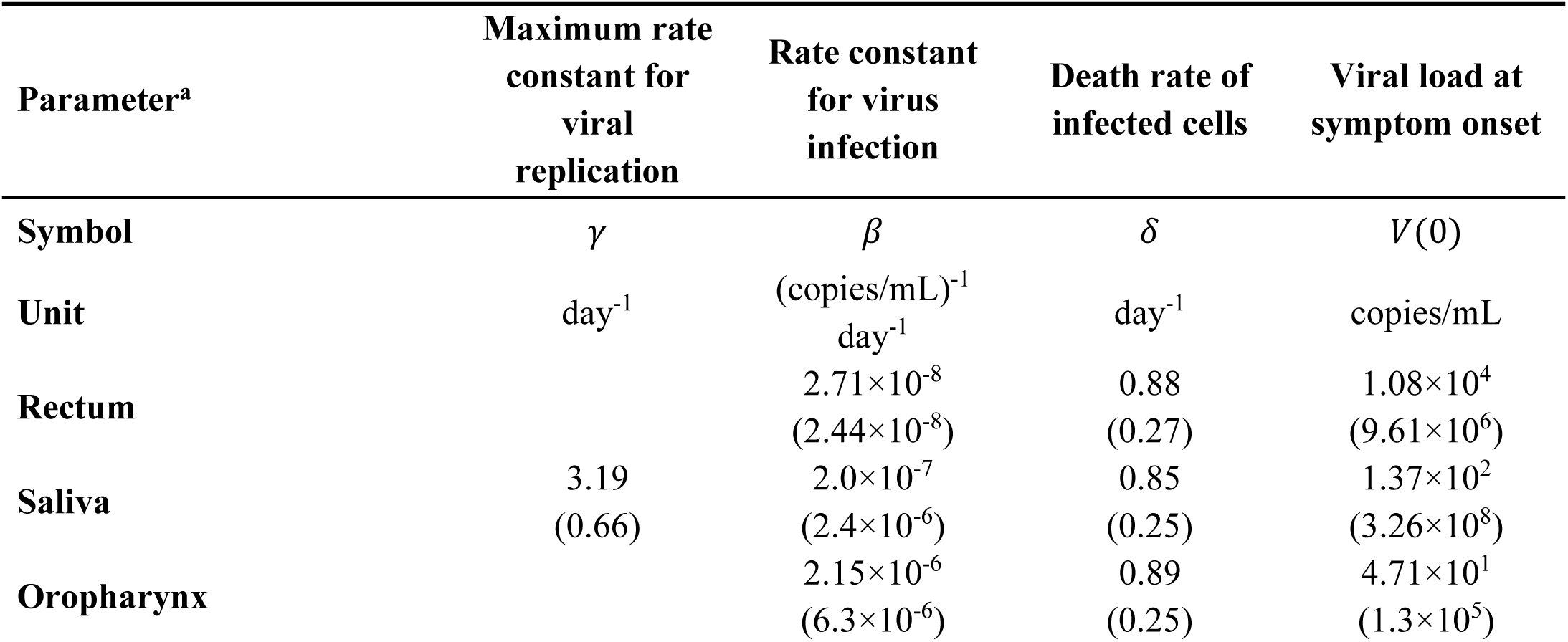
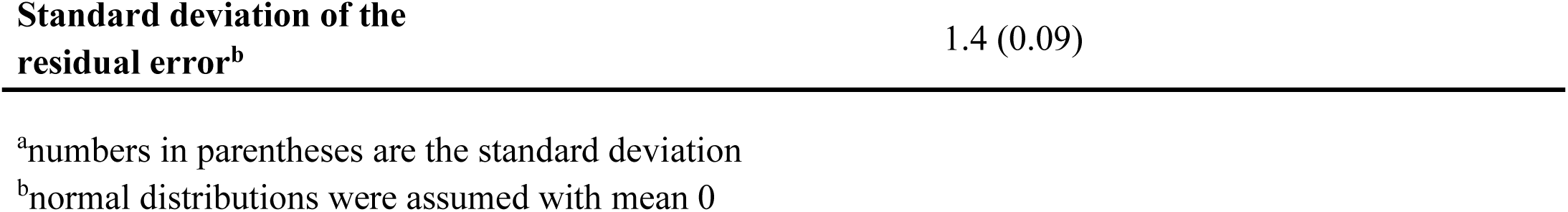
Estimated model parameters for the viral dynamics model.

|  |  |  |  |  |
| --- | --- | --- | --- | --- |
| Parameter<sup>a</sup> | Maximum rate constant for viral replication | Rate constant for virus infection | Death rate of infected cells | Viral load at symptom onset |
| Symbol | $\gamma$ | $\beta$ | $\delta$ | $V(0)$ |
| Unit | day<sup>-1</sup> | (copies/mL)<sup>-1</sup> day<sup>-1</sup> | day<sup>-1</sup> | copies/mL |
| Rectum | | $2.71 \times 10^{-8}$<br>($2.44 \times 10^{-8}$) | 0.88<br>(0.27) | $1.08 \times 10^4$<br>($9.61 \times 10^6$) |
| Saliva | 3.19<br>(0.66) | $2.0 \times 10^{-7}$<br>($2.4 \times 10^{-6}$) | 0.85<br>(0.25) | $1.37 \times 10^2$<br>($3.26 \times 10^8$) |
| Oropharynx | | $2.15 \times 10^{-6}$<br>($6.3 \times 10^{-6}$) | 0.89<br>(0.25) | $4.71 \times 10^1$<br>($1.3 \times 10^5$) |

|  |  |
| --- | --- |
| Standard deviation of the residual error<sup>b</sup> | 1.4 (0.09) |

<sup>a</sup>numbers in parentheses are the standard deviation <sup>b</sup>normal distributions were assumed with mean 0

**Supplemental Table 4.** Mean, median, variance of post-entry incubation period distributions (days)

|  | Mean | Median | Variance |
| --- | --- | --- | --- |
| HS | 5.8 | 4.5 | 23.6 |
| HS+PCR1 | 6.0 | 4.8 | 23.5 |
| HS+PCR2 | 6.0 | 4.8 | 23.5 |
| HS+PCR3 | 6.0 | 4.7 | 23.5 |

Health screening only: HR, Health screening combined with PCR testing at a detection limit of 10 copies/mL: HS+PCR1, 250 copies/mL: HS+PCR2, 1000 copies/mL: HS+PCR3.

## References

1 Van Dijck, C. et al. Emergence of mpox in the post-smallpox era—a narrative review on mpox epidemiology. Clinical Microbiology and Infection 29, 1487–1492 (2023). 10.1016/j.cmi.2023.08.008

2 Bunge, E. M. et al. The changing epidemiology of human monkeypox—A potential threat? A systematic review. PLOS Neglected Tropical Diseases 16, e0010141 (2022). 10.1371/journal.pntd.0010141

3 World Health Organization. Fifth Meeting of the International Health Regulations (2005) (IHR) Emergency Committee on the Multi-Country Outbreak of mpox (monkeypox), <Fifth Meeting of the International Health Regulations (2005) (IHR) Emergency Committee on the Multi-Country Outbreak of mpox (monkeypox)> (

4 Boisson-Walsh, A. Escalating mpox epidemic in DR Congo. The Lancet Infectious Diseases 24, e487 (2024). 10.1016/S1473-3099(24)00446-8

5 Adebisi, Y. A., Ezema, S. M., Bolarinwa, O., Bassey, A. E. & Ogunkola, I. O. Sex Workers and the Mpox Response in Africa. J Infect Dis (2024). 10.1093/infdis/jiae435

6 WHO Director-General declares mpox outbreak a public health emergency of international concern, <https://www.who.int/news/item/14-08-2024-who-director-general-declares-mpox-outbreak-a-public-health-emergency-of-international-concern> (

7 European Centre for Disease Prevention and Control, Communicable disease threats report, 14-20 December 2024, week 51, <https://www.ecdc.europa.eu/en/publications-data/communicable-disease-threats-report-14-20-december-2024-week-51> (

8 Grépin, K. A., Aston, J. & Burns, J. Effectiveness of international border control measures during the COVID-19 pandemic: a narrative synthesis of published systematic reviews. Philos Trans A Math Phys Eng Sci 381, 20230134 (2023). 10.1098/rsta.2023.0134

9 Ponce, L. et al. Incubation Period and Serial Interval of Mpox in 2022 Global Outbreak Compared with Historical Estimates. Emerg Infect Dis 30, 1173–1181 (2024). 10.3201/eid3006.231095

10 Quilty, B. J. et al. Quarantine and testing strategies in contact tracing for SARS-CoV-2: a modelling study. The Lancet Public Health 6, e175–e183 (2021). 10.1016/S2468-2667(20)30308-X

11 Wells, C. R. et al. Optimal COVID-19 quarantine and testing strategies. Nature communications 12, 356 (2021). 10.1038/s41467-020-20742-8

12 Jin, Y., Sun, T., Zheng, P. & An, J. Mass quarantine and mental health during COVID-19: A meta-analysis. Journal of Affective Disorders 295, 1335–1346 (2021). 10.1016/j.jad.2021.08.067

13 World Health Organization. COVID-19: quarantine and self-monitoring, <https://www.who.int/westernpacific/emergencies/covid-19/information/quarantine-and-self-monitoring> (

14 Infection Control Guidance: SARS-CoV-2, <https://www.cdc.gov/covid/hcp/infection-control/index.html> (

15 Lauer, S. A. et al. The Incubation Period of Coronavirus Disease 2019 (COVID-19) From Publicly Reported Confirmed Cases: Estimation and Application. Annals of internal medicine 172, 577–582 (2020). 10.7326/m20-0504

16 Ghani, A. et al. The Early Transmission Dynamics of H1N1pdm Influenza in the United Kingdom. PLoS Curr 1, Rrn1130 (2009). 10.1371/currents.RRN1130

17 Nishiura, H. & Inaba, H. Estimation of the incubation period of influenza A (H1N1-2009) among imported cases: Addressing censoring using outbreak data at the origin of importation. Journal of theoretical biology 272, 123–130 (2011). 10.1016/j.jtbi.2010.12.017

18 Ejima, K. et al. Time variation in the probability of failing to detect a case of polymerase chain reaction testing for SARS-CoV-2 as estimated from a viral dynamics model. J R Soc Interface 18, 20200947 (2021). 10.1098/rsif.2020.0947

19 Yang, Y. et al. Longitudinal viral shedding and antibody response characteristics of men with acute infection of monkeypox virus: a prospective cohort study. Nature communications 15, 4488 (2024). 10.1038/s41467-024-48754-8

20 Brosius, I. et al. Presymptomatic viral shedding in high-risk mpox contacts: A prospective cohort study. Journal of Medical Virology 95, e28769 (2023). 10.1002/jmv.28769

21 Ikeda, H. et al. Quantifying the effect of Vpu on the promotion of HIV-1 replication in the humanized mouse model. Retrovirology 13, 23 (2016). 10.1186/s12977-016-0252-2

22 Martyushev, A., Nakaoka, S., Sato, K., Noda, T. & Iwami, S. Modelling Ebola virus dynamics: Implications for therapy. Antiviral Res 135, 62–73 (2016). 10.1016/j.antiviral.2016.10.004

23 Nowak, M. A. & May, R. M. Virus dynamics : mathematical principles of immunology and virology. (Oxford University Press, 2000).

24 Jeong, Y. D. et al. Revisiting the guidelines for ending isolation for COVID-19 patients. eLife 10, e69340 (2021). 10.7554/eLife.69340

25 Jeong, Y. D. et al. Designing isolation guidelines for COVID-19 patients with rapid antigen tests. Nat Commun 13, 4910 (2022). 10.1038/s41467-022-32663-9

26 Allan-Blitz, L.-T. et al. Laboratory validation and clinical performance of a saliva-based test for monkeypox virus. Journal of Medical Virology 95, e28191 (2023). 10.1002/jmv.28191

27 Nishiura, H. Determination of the appropriate quarantine period following smallpox exposure: An objective approach using the incubation period distribution. International Journal of Hygiene and Environmental Health 212, 97–104 (2009). 10.1016/j.ijheh.2007.10.003

28 Kwok, K. O., Wei, W. I., Tang, A., Wong, S. Y. S. & Tang, J. W. Estimation of local transmissibility in the early phase of monkeypox epidemic in 2022. Clin Microbiol Infect 28, 1653.e1651–1653.e1653 (2022). 10.1016/j.cmi.2022.06.025

29 Suñer, C. et al. Viral dynamics in patients with monkeypox infection: a prospective cohort study in Spain. The Lancet. Infectious diseases 23, 445–453 (2023). 10.1016/s1473-3099(22)00794-0

30 Ministry of Helath, Singapore, Update On Public Health Preparedness Measures for Mpox Clade I, <https://www.moh.gov.sg/news-highlights/details/update-on-public-health-preparedness-measures-for-mpox-clade-i> (

31 De Baetselier, I. et al. Retrospective detection of asymptomatic monkeypox virus infections among male sexual health clinic attendees in Belgium. Nature medicine 28, 2288–2292 (2022). 10.1038/s41591-022-02004-w

32 Agustí, C. et al. Self-sampling monkeypox virus testing in high-risk populations, asymptomatic or with unrecognized Mpox, in Spain. Nature communications 14, 5998 (2023). 10.1038/s41467-023-40490-9

33 Ferré, V. M. et al. Detection of Monkeypox Virus in Anorectal Swabs From Asymptomatic Men Who Have Sex With Men in a Sexually Transmitted Infection Screening Program in Paris, France. Annals of internal medicine 175, 1491–1492 (2022). 10.7326/m22-2183

34 Sabat, J. et al. A comparison of SARS-CoV-2 rapid antigen testing with realtime RT-PCR among symptomatic and asymptomatic individuals. BMC Infect Dis 23, 87 (2023). 10.1186/s12879-022-07969-0

35 Murayama, H. et al. Roles of community and sexual contacts as drivers of clade I mpox outbreaks. medRxiv, 2024.2010.2015.24315554 (2024). 10.1101/2024.10.15.24315554

